# Motor neuron disease in rural Australia: a population-based observational study of epidemiology, clinical characteristics and regional variation

**DOI:** 10.64898/2026.09.08.26362473

**Authors:** Amanda L. Wright, Zoe N. Zussa, Sandrine Chan Moi Fat, Carol M.Y. Lee, Louis Christie, Catherine Keniry, Simon Hawke, Dominic B. Rowe, Kelly L. Williams, Lyndal Henden

## Abstract

**Objectives:** To characterise the epidemiology, clinical features, and geographic distribution of motor neuron disease (MND) within a rural Australian population.

**Design:** Retrospective, population-based epidemiology study.

**Setting:** Western New South Wales Local Health District (WNSWLHD), Australia, from 1 January 2023 to 31 December 2025.

**Participants:** Fifty-one individuals diagnosed with MND identified through Central West Neurology and Neurosurgery, Orange Palliative Care Service, and the WNSWLHD Neurodegenerative Community Care Team.

**Main outcome measures:** Crude and age-standardised incidence, prevalence and mortality rates of MND; demographic and clinical characteristics; geographic distribution of prevalence.

**Results:** The all-age crude incidence of MND was 4.01 per 100,000 person-years (95% CI, 2.78–5.60), prevalence was 8.14 per 100,000 population (95% CI, 6.33–10.30), and mortality was 3.77 per 100,000 person-years (95% CI, 2.58–5.33). Females comprised 51% of patients and showed higher incidence, prevalence and mortality than males. Bulbar-onset MND accounted for 47% of patients, including 61% of females. MND prevalence varied across the health district, with the highest burden observed in central and south-eastern regional areas. Patients travelled on average 139 km to access neurological care, with a diagnostic delay of nine months.

**Conclusions:** MND incidence and mortality are high in this rural Australian population compared with national and global estimates, with an overrepresentation of bulbar-onset disease and altered sex distribution. These findings provide an important baseline for population-level MND surveillance, following the introduction of mandatory notification in NSW, Australia. Findings also highlight the need for strengthened rural healthcare services, and further research of potential genetic and environmental contributors to MND risk.

**Strengths and limitations of this study:**

- This population-based study used multiple clinical and health-service data sources across Western New South Wales Local Health District, with records cross-referenced to maximise case ascertainment and remove duplicates.
- The 2023-2025 study period provides a pre-notification epidemiological baseline preceding the introduction of mandatory motor neuron disease (MND) notification in New South Wales in September 2026.
- Epidemiological and geographical analyses used standardised population denominators and established Australian classifications of remoteness and land use.
- The relatively small number of people with MND limited statistical precision, particularly for subgroup and Local Government Area-level analyses.
- Individual-level genetic, occupational and environmental exposure data were unavailable, limiting investigation of factors potentially underlying demographic and geographic variation.

## Introduction

Motor neuron disease (MND), also known as amyotrophic lateral sclerosis (ALS), is a late-onset neurodegenerative disorder marked by progressive degeneration of motor neurons, leading to paralysis and fatal respiratory failure [1]. MND is more prevalent in males, who typically experience earlier disease onset, whereas females often have a worse prognosis [2–4]. The global burden of MND is increasing [5–7] and in 2021, the age-standardised prevalence and incidence were estimated at 3.31 per 100,000 population and 0.77 per 100,000 person-years, respectively [8], though substantial geographic variation exists between countries [9].

Australia provides a unique setting to examine geographic variation in MND because of its highly polarised population distribution. While major cities occupy less than 1% of the national landmass, almost 70% of the population reside within capital city regions [10]. In contrast, regional and remote Australia spans most of the country and is characterised by sparse populations, large geographic distances, and distinct environmental and occupational exposures. These differences may influence both disease risk and healthcare access.

The importance of examining geographic variation is further supported by emerging evidence that MND may arise from complex interactions between genetic susceptibility and environmental exposures [11]. While 5%–10% of MND are familial and show inheritance of the disease [12], most MND is sporadic and likely multifactorial [13]. Proposed environmental risk factors include agricultural pesticides [14–15], cyanobacterial toxins in waterways [16–17], lead exposure [18–19], and agricultural occupation, particularly crop and livestock farming [20–22]. These observations raise the possibility that rural populations may experience different patterns of disease burden than urban populations.

Contemporary epidemiological data describing the burden of MND in Australia remain limited [23], particularly outside major city centres. While the opt-in MiNDAUS Registry collects national clinical data from people with MND, participation is voluntary and population-level case ascertainment remains limited [24]. A recent Australian study utilising national census data, suggested mortality rates are higher in regional areas than major cities [25], while a population-based study from South Australia reported prevalence estimates exceeding many international cohorts [26]. Fragmented healthcare records and reliance of single-source datasets continue to limit accurate epidemiological assessment, particularly in rural populations. We therefore aimed to characterise the epidemiology, clinical features, and geographic distribution of MND within the Western New South Wales Local Health District (WNSWLHD), the largest rural health district in New South Wales, Australia.

## Material and Methods

The study aimed to identify all people with a confirmed diagnosis of MND residing within WNSWLHD during the period 1 January 2023 to 31 December 2025. Physicians and clinical services across WNSWLHD who frequently diagnose and manage people living with MND were identified through primary site visits. Data collection sites included Central West Neurology and Neurosurgery (Orange, NSW), Orange Palliative Care Service, and the WNSWLHD Neurodegenerative Community Care Team. MND cases were defined as people diagnosed by a neurologist within Australia according to the El Escorial criteria [27]. Clinical and demographic variables were extracted, including date of birth, sex, date of diagnosis, date of symptom onset (where available), date of death where applicable, site of disease onset (bulbar, limb or respiratory), postcode of residence at diagnosis, and the address of the diagnosing neurologist’s clinic and follow-up appointment locations. Information on inheritance classification, genetic aetiology and MND subtype was unavailable for most participants and was excluded from analysis. The consolidated dataset was cross-referenced across all participating collection centres to remove duplicate records for individuals who had been managed by more than one service. A total of 51 people with MND were identified within the WNSWLHD over the three-year study period.

Statistical analyses were conducted in RStudio using R version 4.5.2 [28]. Incidence (number of new MND cases) and mortality (number of MND deaths) rates were calculated annually and as average annual rates across the study period by summing events and dividing by the total person-years at risk. Point prevalence was calculated at December 31 of each year (2023-2025) as the number of people with MND who were alive on that date. Mean annual point prevalence was calculated by summing the revalent cases across the three annual time points and dividing by three times the annual population denominator. Crude incidence and mortality rates were expressed per 100,000 person-years, and prevalence per 100,000 population, with 95% confidence intervals (CIs) assuming a Poisson distribution. Denominator population estimates for WNSWLHD, including for Local Government Areas (LGAs; administrative regions used for local governance and population reporting), were obtained from the 2021 Australian Census. Age-standardised rates were computed by direct standardisation to the 2021 Australian Census population. Ninety-five percent CIs for age-standardised rates were calculated usingByar’s approximation with Dobson’s method adjustment, implemented in the *PHEindicatormethods* R package. Rates were estimated for the total population (all-ages) and for adults aged ≥20, reflecting the adult-onset nature of MND.

Variables derived from clinical records include age at disease onset, duration of disease from onset (until death or last known date of survival) and time from symptom onset to diagnosis (diagnostic delay). The shortest distance travelled by road from residential address to diagnosing clinic address or follow-up appointments was calculated using the Open Source Routing Machine [29]. Since only one individual had respiratory-onset MND, statistical analyses of site of onset were restricted to bulbar and limb onset. A Mann-Whitney U test assessed differences in age at disease onset between both sex and site of onset, between diagnostic delay and site of onset, and between distance travelled to a neurologist and first- or follow-up appointment. Disease duration was evaluated using Kaplan–Meier survival analysis, with individuals who remained alive censored at their last known date of follow-up. Survival distributions were compared by sex and site of onset using log-rank tests. Cox proportional hazards regression was additionally used to estimate hazard ratios (HRs) and 95% confidence intervals (CIs) for mortality according to sex and site of onset. All tests were two-sided, with *P* < 0.05 considered statistically significant.

Maps of population distribution and MND prevalence across LGAs within WNSWLHD were generated using the R packages *nswgeo*, *sf* and *ggplot2*. Remoteness was classified using the Accessibility/Remoteness Index of Australia (ARIA+), as defined within the Australian Statistical Geography Standard (ASGS) remoteness framework [30]. Geographic areas are grouped into five categories according to relative access to services: major cities, inner regional, outer regional, remote or very remote Australia. Rural Australia comprises both regional and remote areas. Land use is specified according to the Australian Land Use and Management (ALUM) Classification version 8 [31] using the simplified classification system. Spatial data were obtained from the NSW Land Use 2017 dataset (v1.5) [32].

### Ethics approval

Human research ethics was approved by the Greater Western Human Research Ethics Committee (HREC; 2025/ETH00163) and the Charles Sturt University HREC (H25318) in accordance with the Declaration of Helsinki. The requirement for informed consent was waived by the approving committees.

## Results

### Western NSW Local Health District is geographically vast with a low and dispersed population density

WNSWLHD covers approximately 247,000 km² (Figure 1), encompassing 30.7% of New South Wales and 3.2% of Australia’s landmass. The district supports a population of 283,564 people, representing 1.1% of the Australian population (2021 census). For global context across continents, the land area of this single health district is comparable in size to the United Kingdom (Europe, 67 million population), Oregon state (North America, 4.2 million population), Ecuador (South America, 18.3 million population), Laos (Asia, 8 million population) and Uganda (Africa, 48.9 million population), but with a substantially smaller population. The underlying WNSWLHD population demonstrated a male-to-female ratio of 1:1 overall, with a shift towards female predominance for people aged ≥80.

**Figure 1.**
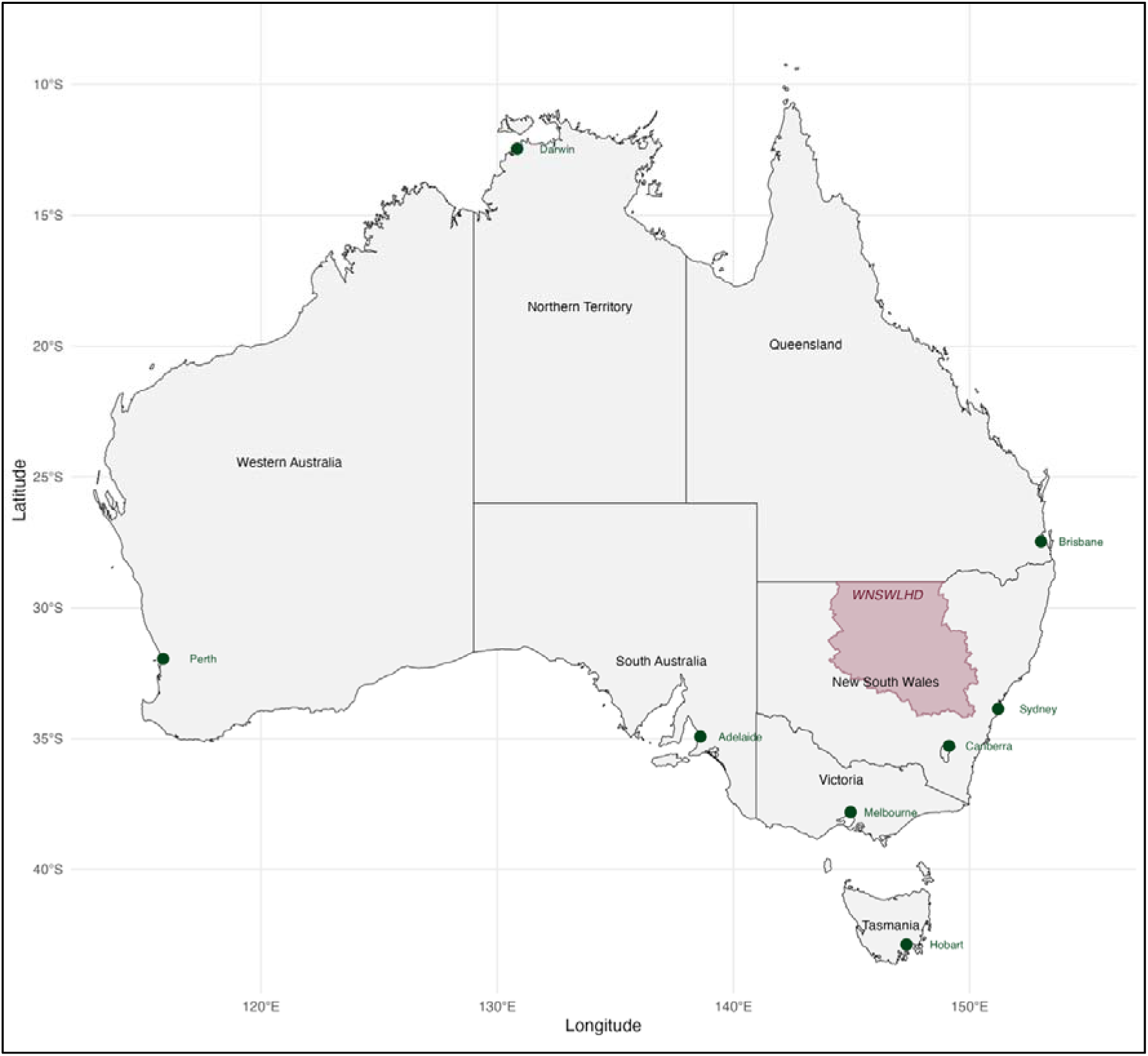
Geographic location of WNSWLHD highlighted in pink on a map of Australia, indicating the source of collected data. Major cities are denoted by green dots.

The WNSWLHD region is divided into 22 LGAs (Figure 2A). Population density is highest in the south-east LGAs, particularly Dubbo, Orange and Bathurst. In contrast, LGAs located further west (inland Australia) have progressively smaller and more dispersed populations. This population distribution aligns with the Australian Bureau of Statistics remoteness classifications, which reflect relative geographic access to services. Approximately 70% of WNSWLHD is classified as remote or very remote, while the remaining 30% is categorised as inner or outer regional Australia (Figure 2B). No areas within WNSWLHD are classified as major cities.

**Figure 2.**
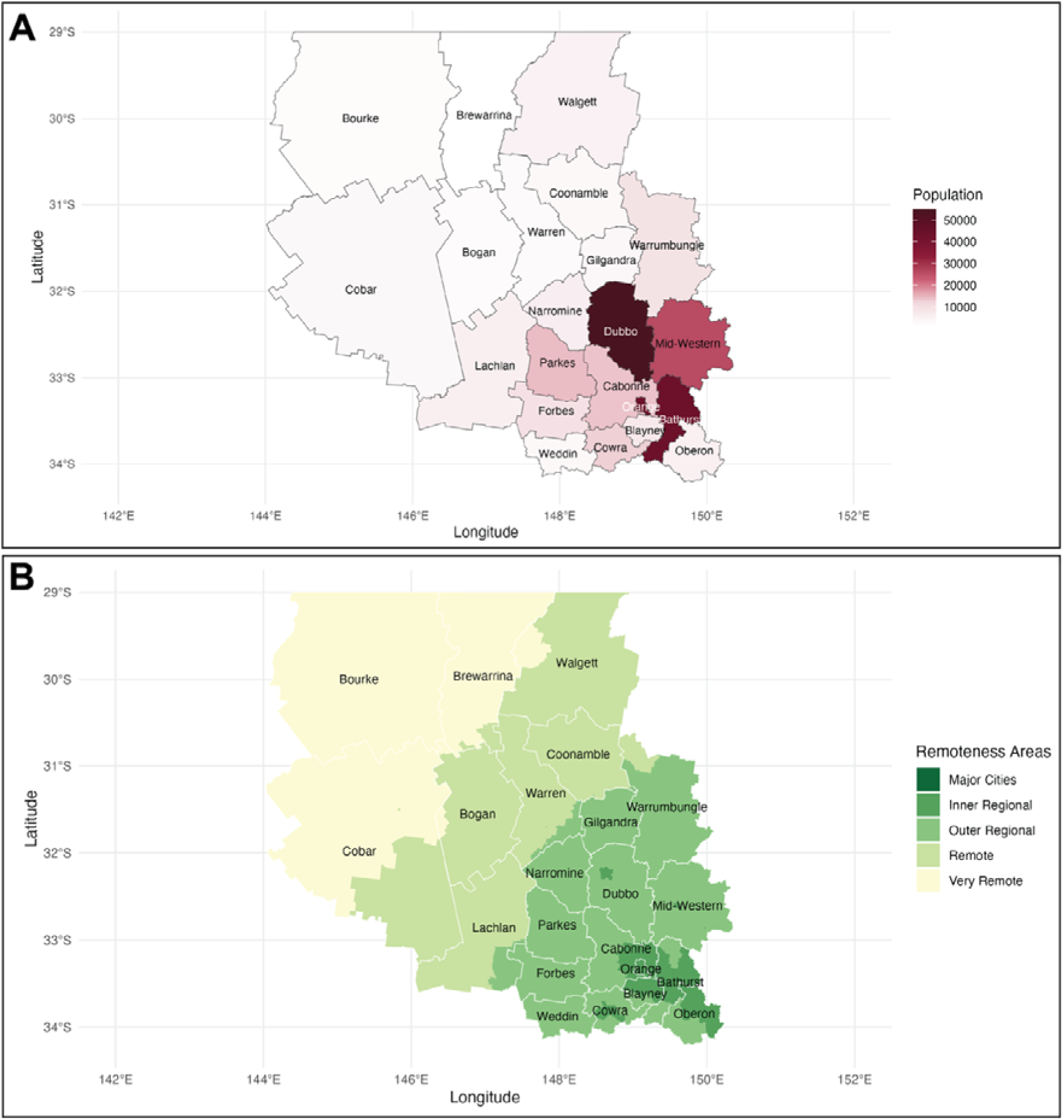
(A) Map outlining the Local Government Areas (LGAs) within WNSWLHD, showing population distribution. **(B)** Remoteness areas within WNSWLHD, classified according to the Australian Bureau of Statistics remoteness structure.

Land use across WNSWLHD is predominantly agricultural and varies by region (Figure 3). Almost 60% of land, primarily the north-west, is used for grazing native vegetation by domestic livestock (cattle and sheep), with minimal pasture modification. Broadacre dryland and irrigated cropping account for a further 21% of land use, concentrated in central regions and supporting crops such as wheat, barley, hay, cotton and canola (oilseed rape) [31]. Grazing livestock on modified pastures, including fertilised or introduced species to support high-production livestock systems, represents around 7% of land use and is more common in the eastern areas of the district.

**Figure 3.**
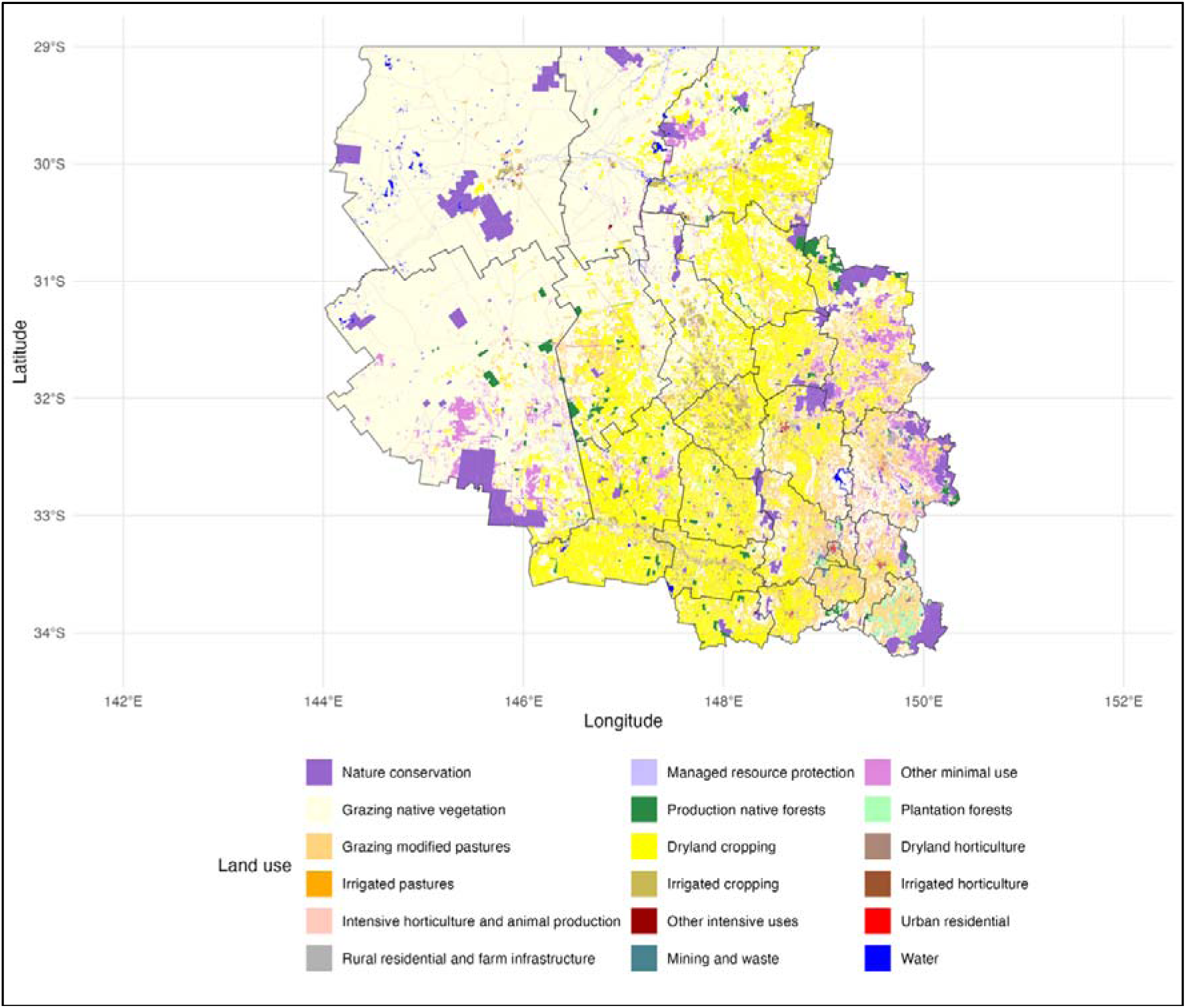
Land use within WNSWLHD. Land use categories are defined according to the Australian Land Use and Management (ALUM) Classification Version 8 using the simplified classification symbology.

### Equal male-to-female ratio and bulbar-onset enrichment in WNSWLHD

Between 1 January 2023 to 31 December 2025, a total of 51 people currently living or newly diagnosed with MND were identified within WNSWLHD using medical records and databases from three collection services. To the best of our knowledge, ascertainment through these sources captures all MND cases residing within WNSWLHD during the study period. Demographic and clinical characteristics are summarised in Table 1. Of these, 51% were female (Figure 4A). The average age at disease onset was higher in females (69.9 years) than in males (61.7 years), although this difference did not reach statistical significance (Figure 4B, *P* = 0.33). Median survival was shorter in females compared to males (25.6 vs 57.6 months, Figure 4C, *P* = 0.047), with females having an estimated 2.81-fold higher hazard of death (HR 2.81, 95% CI 0.98–8.06). Limb-onset disease was the most common presentation (51%), though presentation with bulbar-onset disease was high (47%), while respiratory-onset MND was rare (2%) (Figure 4D). Individuals with bulbar-onset MND had disease onset approximately 10-years later than those with limb-onset (70.1 vs 60.9 years, Figure 4E, *P* = 0.044). Median survival was shorter in individuals with bulbar-onset disease than limb-onset disease (23.7 vs 52.8 months, Figure 4F, *P* = 0.026), corresponding to a 2.92-fold higher hazard of death (HR 2.92, 95% CI 1.09–7.83). Bulbar-onset MND was more common among females, with 61% of females presenting with bulbar-onset disease compared to only 33% of males, though this difference was not statistically significant (Figure 4G, *P* = 0.14). The average time from symptom onset to diagnosis was nine months and did not differ between bulbar- and limb-onset disease (Figure 4H, *P* = 0.66). Patients travelled an average distance of 139 km (86 miles) to attend their initial neurology consultation. Follow-up appointments were conducted at greater distances, although this difference was minimal and not statistically significant (median difference of 7 km, Figure 4I, *P* = 0.99).

**Figure 4.**
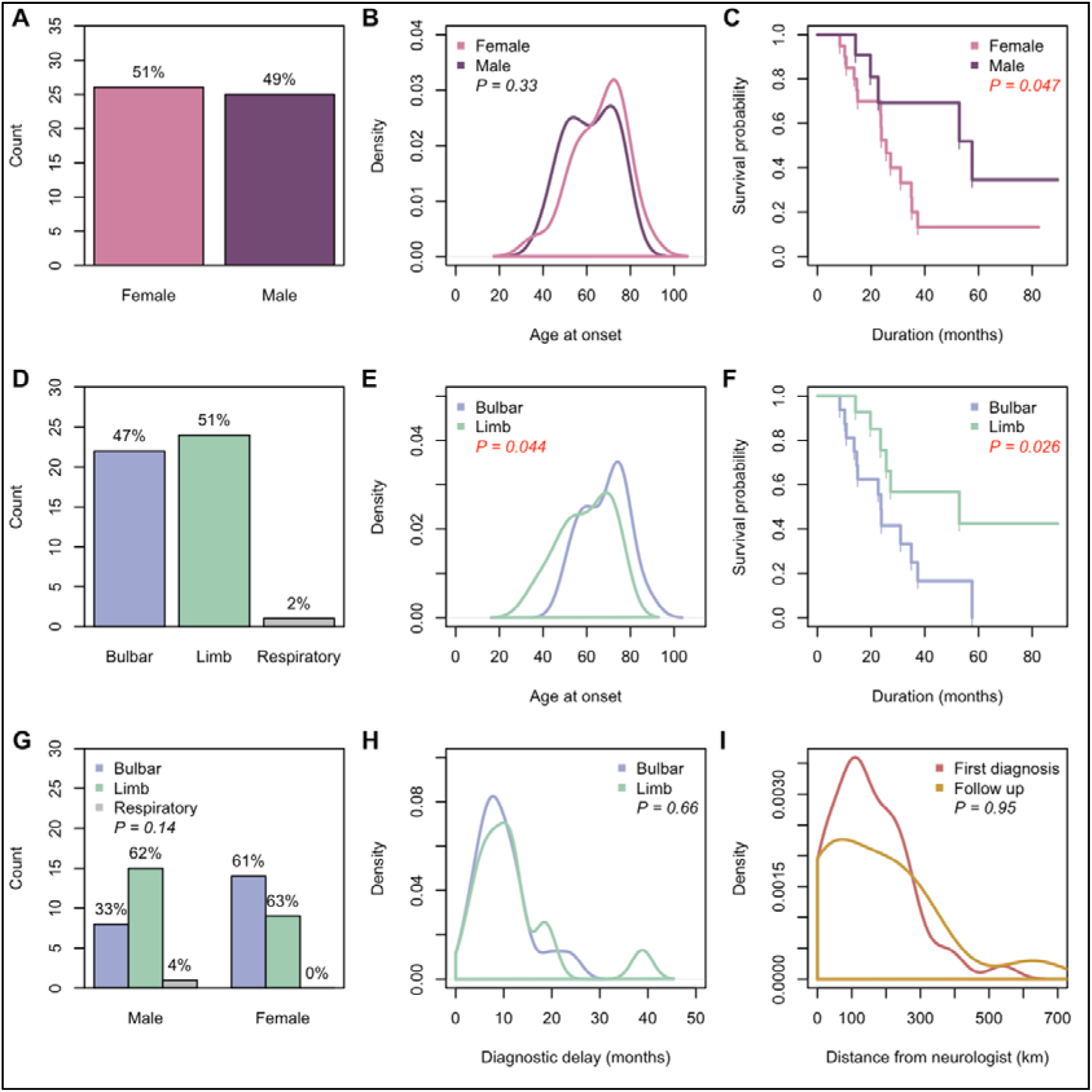
Demographic and clinical characteristics of the rural Australian MND cohort. Overview of key demographic features, disease onset patterns, and survival outcomes within the cohort. **(A)** Sex distribution demonstrating a higher proportion of female to males diagnosed with MND. **(B)** Age at symptom onset revealed no sex differences. **(C)** Kaplan– Meier survival analysis comparing males and females revealed a significant reduction in survival of females. **(D)** Distribution of disease onset types across the cohort. **(E)** Age of onset according to site of disease onset (bulbar vs limb) showed a significant early onset of limb onset phenotype. **(F)** Survival analysis by site of onset, highlighting a significant difference in disease progression between bulbar and limb onset phenotypes. **(G)** Distribution of males and females by site of onset showed a higher proportion of females with bulbar onset. **(H)** Diagnostic delay according to site of onset, illustrating no differences in time from symptom onset to diagnosis across limb and bulbar subtypes. **(I)** Travel distance from first and follow up neurology appointment shows no difference in travel distances.

**Table 1.** Demographic and clinical characteristics of the WNSWLHD MND cohort. Age at disease onset, diagnostic delay and distance travelled to a neurologist are presented as median (interquartile range). Disease duration from onset is presented as Kaplan–Meier median survival (95% CI), accounting for censoring at the last known date of survival. NE indicates that the upper confidence limit was not estimable within the observed follow-up period. Respiratory-onset MND was excluded from site of onset comparisons due to the small number of cases (*n*=1).

| **Sex** |  |  |
| --- | --- | --- |
|  | *Male* | *Female* |
| *n* (%) | 25 (49%) | 26 (51%) |
| Age at disease onset (years) | 61.7 (50.6 – 71.9) | 69.9 (57.5 – 75.1) |
| Kaplan-Meier median survival (months) | 57.6 (22.6 – NE) | 25.6 (23.5 – 37.4) |
| **Site of onset** |  |  |
|  | *Limb* | *Bulbar* |
| *n* (%) | 24 (51%) | 22 (47%) |
| Age at disease onset (years) | 60.9 (51.0 – 70.6) | 70.1 (60.3 – 75.5) |
| Kaplan-Meier median survival (months) | 52.8 (25.6 – NE) | 23.7 (14.6 – NE) |
| Diagnostic delay (months) | 10.0 (6.5 – 12.0) | 8.7 (6.0 – 12.0) |
| **Distance to neurologist** |  |  |
|  | *First appointment* | *Follow up appointment* |
| Distance travelled by road (km) | 139.3 (89.8 – 223.5) | 147.5 (28.4 – 261.3) |

### Incidence, prevalence and mortality of MND in WNSWLHD, with higher rates in females

Average annual incidence, prevalence and mortality rates are provided in Table 2. Across the three-year study period, the average annual all-age crude incidence rate in WNSWLHD was 4.01 per 100,000 person-years (95% CI, 2.78–5.60). The corresponding crude prevalence was 8.14 per 100,000 population (95% CI, 6.33–10.30) and crude mortality was 3.77 per 100,000 person-years (95% CI, 2.58–5.33). Age-standardised rates were lower, reflecting the older age demographic of WNSWLHD compared to the Australian population, with an incidence of 3.72 per 100,000 person-years (95% CI, 2.57–5.20), prevalence of 7.50 per 100,000 population (95% CI, 5.83–9.50) and mortality of 3.40 per 100,000 person-years (95% CI, 2.33–4.81). Restricting analyses to adults aged ≥ 20 years (Table 2) increased incidence to 4.89 per 100,000 person-years (95% CI, 3.38–6.84), increased prevalence to 9.86 per 100,000 population (95% CI, 7.67–12.49) and increased mortality to 4.48 per 100,000 person-years (95% CI, 3.06–6.33), an overall increase of 30-36% compared with all-age estimates.

**Table 2.** Incidence, prevalence and mortality of MND in WNSWLHD. Crude rates were calculated using the 2021 Australian Bureau of Statistics census population for WNSWLHD, while age-standardised rates were derived using the Australian population from the 2021 census. Rates are reported for the total population (all-age, 0-85+ years) and for adults aged ≥ 20 years. 95% confidence intervals are reported within brackets.

| **Method** | **Population** | **Incidence**<br>**per 100,000**<br>**person-years** | **Prevalence**<br>**per 100,000**<br>**population** | **Mortality**<br>**per 100,000**<br>**person-years** |
| --- | --- | --- | --- | --- |
| Crude | All-age | 4.01 (2.78 – 5.60) | 8.14 (6.33 – 10.30) | 3.77 (2.58 – 5.33) |
| Crude | $\geq 20$ | 5.44 (3.77 – 7.60) | 11.04 (8.59 – 13.97) | 5.12 (3.50 – 7.23) |
| Age<br>standardised | All-age | 3.72 (2.57 – 5.20) | 7.50 (5.83 – 9.50) | 3.40 (2.33 - 4.81) |
| Age<br>standardised | $\geq 20$ | 4.89 (3.38 - 6.84) | 9.86 (7.67 - 12.49) | 4.48 (3.06 - 6.33) |

Age-specific analyses demonstrated an increase in MND burden with advancing age (Table S1). Incidence, prevalence and mortality were highest in the 75–79-year age group, while cases were rare below 50 years of age. This pattern is consistent with the established age-dependent epidemiology of MND. The estimated net cumulative incidence of MND to age 85 was approximately 1 in 261 individuals, assuming no competing mortality.

Sex-stratified estimates are presented in Table S2. Crude incidence was higher in females than males (4.74 vs 3.29 per 100,000 person-years), corresponding to a female-to-male rate ratio (RR) of 1.44 (95% CI, 0.73–2.86). Similarly, crude prevalence (8.53 vs 7.75 per 100,000 population; RR 1.10, 95% CI, 0.69–1.77) and mortality (4.50 vs 3.05 per 100,000 person-years; RR 1.48, 95% CI, 0.73–2.99) were higher in females. The same pattern was observed for age-standardised estimates, with consistently higher rates in females than males, though wide confidence intervals around the rate ratios all included 1, indicating no statistically significant sex differences.

Annual analyses showed relatively stable incidence rates in recent years, with crude estimates of 4.60 per 100,000 person-years (95% CI, 2.45–7.87) in 2023, 4.95 per 100,000 person-years (95% CI, 2.71–8.31) in 2024, and 2.48 per 100,000 person-years (95% CI, 1.00–5.10) in 2025 (Table S3). Prevalence estimates were 8.85 (95% CI, 5.73–13.06), 9.56 (95% CI, 6.30–13.90), and 6.02 (95% CI, 3.50–9.63) per 100,000 population in 2023, 2024 and 2025, respectively. Mortality rates showed steady increases across all three years, at 1.77 (95% CI, 0.57–4.13), 3.54 (95% CI, 1.70-6.51), and 6.02 (95% CI, 3.50-9.63) per 100,000 person-years in 2023, 2024 and 2025, respectively.

### Geographic variation in MND prevalence across WNSWLHD

MND prevalence varies across the LGAs within WNSWLHD (Figure 5). The highest prevalence estimates were observed in Oberon (23.90 per 100,000 population, 95% CI, 6.51–61.18) and Walgett (19.04 per 100,000 population, 95% CI, 3.93–55.63). Elevated prevalence was also observed in Forbes, Cowra, Orange, and Mid-Western LGAs (6.48–10.70 per 100,000 population). In contrast, no cases were recorded during the study period in several neighbouring LGAs, including Bogan, Cobar, and Weddin. The geographic distribution suggests heterogeneity in MND prevalence across the health district, with higher rates observed in specific regional centres and surrounding communities, particularly in the central and south-eastern regions of WNSWLHD.

**Figure 5.**
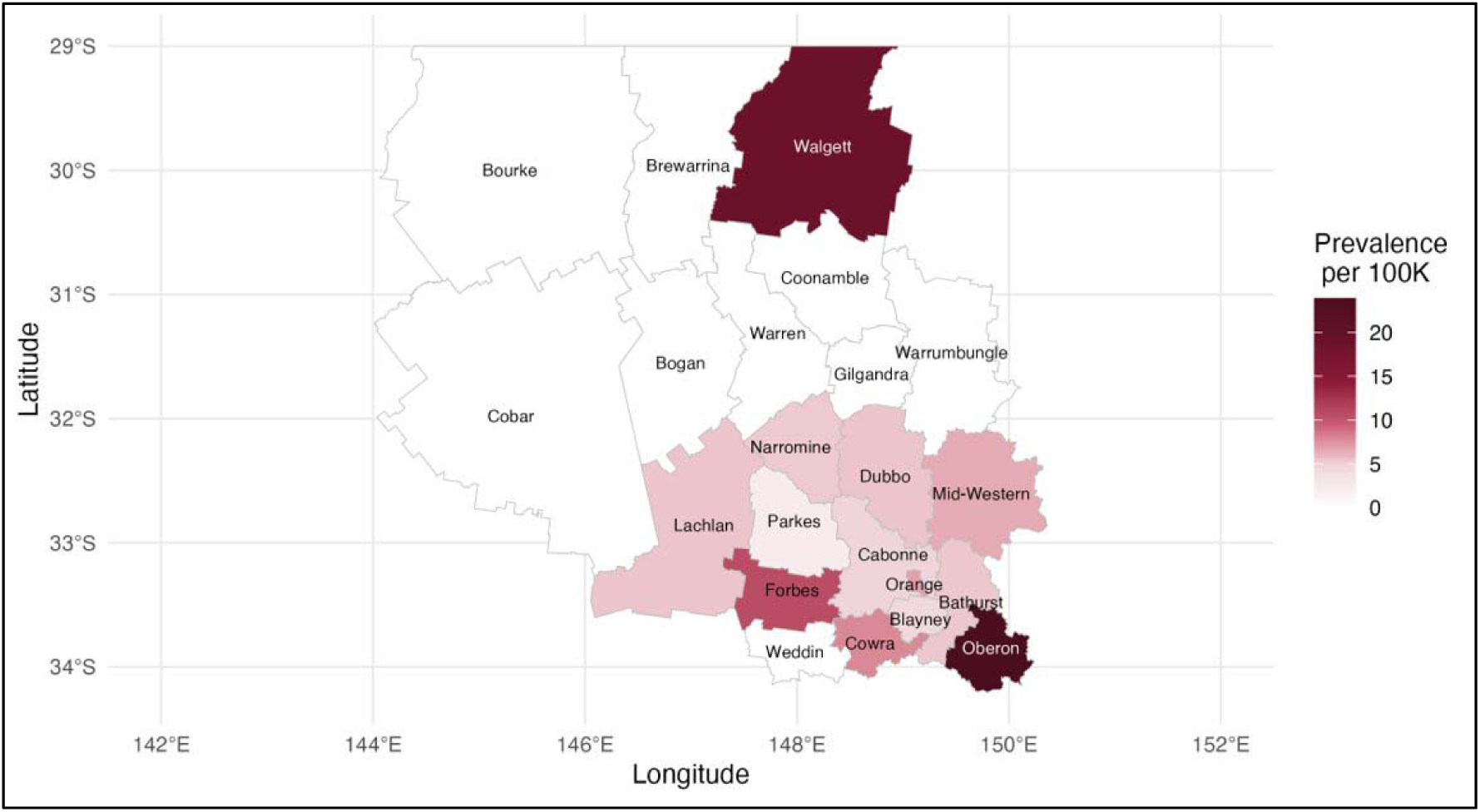
Geographic distribution of MND prevalence across Local Government Areas (LGAs) within WNSWLHD. MND prevalence rates were calculated using LGA population estimates from the 2021 Australian Bureau of Statistics census and mapped to illustrate geographic variation across the region.

## Discussion

This study provides the first comprehensive epidemiological and clinical characterisation of MND within a rural Australian population, identifying a distinct regional clinical profile and increased burden of disease within WNSWLHD that warrants further investigation.

A higher proportion of bulbar-onset MND was observed in WNSWLHD than typically reported within Australia or globally. Nearly half of patients (47%) presented with bulbar-onset disease, compared with the expected 20–30% [1, 4, 33]. Previous population-based analyses have demonstrated that females are more likely to present with bulbar-onset disease than males [2]. While MND commonly demonstrates male predominance (∼1.25:1 [34]), the near 1:1 male-to-female ratio observed in WNSWLHD was notable, though did not fully explain the elevated proportion of bulbar-onset disease. Instead, the site-of-onset distribution among females appeared shifted, with 61% presenting with bulbar-onset MND compared with the 30–40% generally reported [4, 33]. The drivers of these distinct demographic and clinical features are likely multifactorial. The WNSWLHD population is older than the national average, and increasing age is associated with a higher likelihood of bulbar-onset disease. Ascertainment bias may also contribute, particularly in settings where individuals with more rapidly progressive or clinically apparent disease are more likely to be diagnosed. However, broader shifts in sex distribution have recently been reported elsewhere and are attributed to improved case ascertainment and increasing female participation in occupations and environmental settings associated with higher MND risk [35]. Genetic factors represent another possible explanation, as sex ratios appear more balanced in genetically determined disease [34]. However, genetic data was unavailable as genetic diagnostic screening is not currently routinely performed, potentially obscuring underlying genetic contributions within this population.

The burden of MND observed in WNSWLHD was high relative to both Australian and international estimates. The all-age crude incidence of 4.01 per 100,000 person-years exceeded that reported in the only comparable Australian population-based study from South Australia (3.34 per 100,000 person-years) [26], which is itself considered high by international standards. Incidence estimates were comparable to those reported in England, Ireland, Scotland and Sweden (3.83–5.75 per 100,000 person-years) [36–39], countries recognised as having among the highest reported MND incidence globally [9]. Mortality within WNSWLHD was similarly elevated, with an all-age crude mortality of 3.77 per 100,000 person-years compared with the recent Australian national estimate of 2.93 per 100,000 person-years [25]. Mortality estimates in WNSWLHD were also consistent with the increased mortality observed in inner and outer regional Australia (3.24–3.90 per 100,000 person-years) [25]. Importantly, this pattern persisted after age standardisation, indicating that the increased burden was not solely attributable to the older demographic structure of WNSWLHD.

Despite elevated incidence, prevalence within WNSWLHD was not proportionally increased. This likely reflects the clinical composition of the cohort, characterised by enrichment of bulbar-onset disease and higher representation of females, both associated with poorer prognosis and shorter survival [4, 33]. Consequently, increased incidence may not translate into proportionally higher prevalence, as individuals spend less time living with the disease. This suggest that rural populations may experience a distinct pattern of MND burden characterised by high incidence and mortality but comparatively shorter disease duration.

Geographic variation in MND prevalence was observed across WNSWLHD. Higher prevalence occured within central and south-eastern LGAs, corresponding to areas with greater population density and predominantly inner and outer regional classifications. In contrast, no cases were recorded in several sparsely populated remote western LGAs. This pattern aligns with recent Australian mortality data demonstrating higher MND burden in inner and outer regional areas compared with remote regions [25]. Geographic variation also overlapped with differences in land use across the district. Regions with higher prevalence coincided with regions characterised by more intensive agricultural activity, including cropping and modified pasture systems. This observation is consistent with reports of increased prevalence of bulbar-onset MND among agricultural workers [40]. Although causal inference cannot be established, these findings support the need for further investigation into potential environmental and occupational contributors to disease risk in rural Australia. However, interpretation must consider the substantial geographic and healthcare disparities across WNSWLHD, where approximately 70% of the district is classified as remote or very remote. Differences in healthcare access, diagnostic pathways, and population density are likely to influence observed prevalence patterns.

Interestingly, diagnostic delay within WNSWLHD was shorter than reported in previous Australian studies [41]. Although patients travelled substantial distances to access neurological care (mean 139 km), the average diagnostic delay was approximately nine months compared with around 13 months nationally [41]. These findings suggest that timely diagnosis can still be achieved within regional healthcare systems despite significant geographic barriers.

This study has several limitations. The modest sample size limited statistical power for subgroup and regional analyses, resulting in wide and overlapping confidence intervals. Accordingly, observed differences, including sex-specific rate estimates and variation between LGAs, should be interpreted as descriptive patterns rather than definitive differences. This is particularly relevant for LGAs with small resident populations, where small changes in case numbers can substantially affect prevalence estimates. The absence of genetic data and the inability to reliably distinguish familial from sporadic disease restricted assessment of potential genetic contributions to the observed geographic patterns. Future studies incorporating genetic data may provide further insight into these regional differences.

Case ascertainment relied on multiple regional clinical and service-based data sources, which strengthened case identification but may not have captured all cases. Population movement into and out of the region could also not be reliably accounted for, reflecting the limitations of retrospective regional surveillance in the absence of a mandatory population-based registry.

## Conclusion

The findings reported here have important implications for both research and healthcare delivery. The magnitude and geographical distribution of MND observed within WNSWLHD suggest that the burden of disease in regional and remote Australia may be underestimated. The introduction of mandatory MND notification in NSW from September 2026, following implementation of the Public Health Amendment (Motor Neurone Disease) Order 2026, provides an important new framework for prospective population-level surveillance. The potential value of mandatory reporting is illustrated by experience in Massachusetts, USA where only 61.6% of ALS cases captured by the compulsory state registry were also identified in the non-notifiable US National ALS Registry [42]. Prospective notification in NSW will provide an opportunity to evaluate regional incidence and geographic patterns against the pre-notification estimates reported here. More broadly, this work highlights the importance of geographically informed approaches to MND research and the need for strengthened rural neurological services and infrastructure to address the growing burden of MND in Australia.

## Data Availability

Due to the sensitive nature of the data and the small sample size, which increases the risk of participant re-identification, the data are not publicly available.

## Author Contribution

Conceptualisation: Amanda L. Wright, Kelly L. Williams, Lyndal Henden. Resources: Louis Christie, Simon Hawke. Data curation: Amanda L. Wright, Zoe N. Zussa, Sandrine Chan Moi Fat, Lyndal Henden. Formal analysis: Amanda L. Wright, Lyndal Henden. Investigation: Amanda L. Wright, Lyndal Henden. Project administration: Catherine Keniry. Supervision: Dominic B. Rowe, Kelly L. Williams, Lyndal Henden. Writing – original draft: Amanda L. Wright, Lyndal Henden. Writing – review and editing: Amanda L. Wright, Zoe N. Zussa, Sandrine Chan Moi Fat, Carol M.Y Lee, Louis Christie, Catherine Keniry, Simon Hawke, Dominic B. Rowe, Kelly L. Williams, Lyndal Henden. All authors have read and approved the final manuscript.

## Acknowledgements

The authors gratefully acknowledge the support of the Neurodegenerative Community Care Team for their assistance in patient coordination and data access. The authors acknowledge the administration staff of Central West Neurology and Neurosurgery for their assistance in case ascertainment. We also extend our sincere thanks to the patients and their families whose participation made this research possible.

## Funding

ZNZ is supported by MotorOn. KLW is supported by FightMND Bill Guest Mid-Career Research Fellowship and NHMRC Investigator Grant (GNT2033019). DBR and CMYL are supported by the New South Wales Ministry of Health Motor Neuron Disease Research Grant (341925612).

## Artificial Intelligence

During the preparation and revision of this manuscript, the authors used ChatGPT (OpenAI; GPT-5.2) to improve the clarity and readability of content written by the authors. Following use of this tool, the authors reviewed and edited all content as appropriate and take full responsibility for the content of the publication.

## Conflicts of Interests

The authors report there are no competing interests to declare.

## Notes

### Competing Interest Statement

The authors have declared no competing interest.

