## Supplementary Material for "Motor neuron disease in rural Australia: a population-based observational study of epidemiology, clinical characteristics and regional variation"

Amanda Wright *et al.*

**This file includes:**

Supplementary Tables 1 – 3

| Age group | Incidence per 100,000 person-years | | | Prevalence per 100,000 population | | | Mortality per 100,000 person-years | | |
|---|---|---|---|---|---|---|---|---|---|
| | All | Male | Female | All | Male | Female | All | Male | Female |
| 20-24 years | 0 (0 - 8.08) | 0 (0 - 15.58) | 0 (0 - 16.79) | 0 (0 - 8.08) | 0 (0 - 15.58) | 0 (0 - 16.79) | 0 (0 - 8.08) | 0 (0 - 15.58) | 0 (0 - 16.79) |
| 25-29 years | 0 (0 - 6.97) | 0 (0 - 14.01) | 0 (0 - 13.89) | 0 (0 - 6.97) | 0 (0 - 14.01) | 0 (0 - 13.86) | 0 (0 - 6.97) | 0 (0 - 14.01) | 0 (0 - 13.86) |
| 30-34 years | 0 (0 - 6.78) | 0 (0 - 13.55) | 0 (0 - 13.58) | 0 (0 - 6.78) | 0 (0 - 13.55) | 0 (0 - 13.58) | 0 (0 - 6.78) | 0 (0 - 13.55) | 0 (0 - 13.58) |
| 35-39 years | 1.97 (0.05 - 10.99) | 0 (0 - 14.50) | 3.96 (0.10 - 22.04) | 1.97 (0.05 - 10.99) | 0 (0 - 14.50) | 3.96 (0.10 - 22.04) | 0 (0 - 7.27) | 0 (0 - 14.5) | 0 (0 - 14.59) |
| 40-44 years | 2.18 (0.06 - 12.16) | 4.42 (0.11 - 24.62) | 0 (0 - 15.90) | 2.18 (0.06 - 12.16) | 4.42 (0.11 - 24.62) | 0 (0 - 15.90) | 0 (0 - 8.05) | 0 0 - 16.3) | 0 (0 - 15.9) |
| 45-49 years | 0 (0 - 7.45) | 0 (0 - 15.01) | 0 (0 - 14.78) | 2.02 (0.05 - 11.25) | 4.07 (0.10 - 22.67) | 0 (0 - 14.78) | 2.02 (0.05 - 11.25) | 4.07 (0.1 - 22.67) | 0 (0 - 14.78) |
| 50-54 years | 11.41 (4.19 - 24.82) | 11.52 (2.38 - 33.66) | 11.29 (2.33 - 33.00) | 17.11 (7.82 - 32.48) | 3.84 (0.10 - 21.39) | 30.11 (13.00 - 59.34) | 1.9 (0.05 - 10.59) | 0 (0 - 14.16) | 3.76 (0.1 - 20.97) |
| 55-59 years | 3.7 (0.45 - 13.35) | 3.68 (0.09 - 20.52) | 3.71 (0.09 - 20.66) | 14.78 (6.38 - 29.13) | 22.10 (8.11 - 48.11) | 7.41 (0.90 - 26.78) | 5.54 (1.14 - 16.2) | 0 (0 - 13.59) | 11.12 (2.29 - 32.5) |
| 60-64 years | 5.58 (1.15 - 16.32) | 0 (0 - 13.72) | 11.18 (2.30 - 32.66) | 13.03 (5.24 - 26.85) | 3.72 (0.09 - 20.73) | 22.25 (8.20 - 48.65) | 5.58 (1.15 - 16.32) | 3.72 (0.09 - 20.73) | 7.45 (0.9 - 26.92) |
| 65-69 years | 4.24 (0.51 - 15.33) | 8.35 (1.01 - 30.16) | 0 (0 - 15.93) | 23.35 (11.65 - 41.77) | 33.40 (14.42 - 65.80) | 12.95 (2.67 - 37.86) | 14.86 (5.97 - 30.61) | 20.87 (6.78 - 48.71) | 8.64 (1.05 - 31.19) |
| 70-74 years | 18.17 (7.85 - 35.81) | 13.56 (2.80 - 39.62) | 22.84 (7.42 - 53.30) | 24.99 (12.47 - 44.71) | 31.63 (12.72 - 65.17) | 18.27 (4.98 - 46.78) | 13.63 (5 - 29.67) | 9.04 (1.09 - 32.65) | 18.27 (4.98 - 46.78) |
| 75-79 years | 24.98 (10.78 - 49.21) | 18.89 (3.90 - 55.20) | 30.96 (10.05 - 72.25) | 40.59 (21.61 - 69.40) | 18.89 (3.90 - 55.20) | 61.92 (29.69 - 113.88) | 18.73 (6.87 - 40.77) | 12.59 (1.53 - 45.49) | 24.77 (6.75 - 63.42) |
| 80-84 years | 4.54 (0.11 - 25.29) | 0 (0 - 35.51) | 8.59 (0.22 - 47.84) | 22.69 (7.37 - 52.95) | 38.5 (10.49 - 98.58) | 8.59 (0.22 - 47.84) | 13.61 (2.81 - 39.79) | 9.63 (0.24 - 53.63) | 17.17 (2.08 - 62.04) |
| 85+ years | 9.92 (1.20 - 35.85) | 12.91 (0.33 - 71.93) | 8.06 (0.20 - 44.90) | 9.92 (1.20 - 35.85) | 12.91 (0.33 - 71.93) | 8.06 (0.20 - 44.90) | 9.92 (1.2 - 35.85) | 12.91 (0.33 - 71.93) | 8.06 (0.2 - 44.9) |

**Supplementary Table 1.** Incidence, prevalence and mortality of MND in WNSWLHD for each five-year age group, starting at 20 years. Crude rates were calculated using the 2021 Australian Bureau of Statistics census population for WNSWLHD. 95% confidence intervals are included in brackets.

| Method | Population | Incidence per 100,000 person-years | | | Prevalence per 100,000 population | | | Mortality per 100,000 person-years | | |
|---|---|---|---|---|---|---|---|---|---|---|
| | | Male | Female | Risk ratio | Male | Female | Risk ratio | Male | Female | Risk ratio |
| Crude | All-age | 3.29 (1.80 - 5.52) | 4.74 (2.90 - 7.32) | 1.44 (0.73 - 2.86) | 7.75 (5.33 - 10.88) | 8.53 (5.98 - 11.82) | 1.10 (0.69 - 1.77) | 3.05 (1.63 - 5.22) | 4.50 (2.71 - 7.03) | 1.48 (0.73 - 2.99) |
| Age-standardised | All-age | 3.06 (1.66 - 5.15) | 4.40 (2.68 - 6.80) | 1.39 (0.70 - 2.76) | 6.99 (4.80 - 9.83) | 7.98 (5.58 - 11.06) | 1.06 (0.66 - 1.70) | 2.72 (1.44 - 4.66) | 4.1 (2.46 - 6.4) | 1.42 (0.70 - 2.88) |
| Crude | Populations ≥20 | 4.52 (2.47 - 7.58) | 6.35 (3.88 - 9.81) | 1.41 (0.71 - 2.79) | 10.64 (7.33 - 14.95) | 11.43 (8.01 - 15.83) | 1.07 (0.67 - 1.72) | 4.19 (2.23 - 7.17) | 6.04 (3.63 - 9.42) | 1.44 (0.71 - 2.91) |
| Age-standardised | Populations ≥20 | 4.08 (2.22 - 6.87) | 5.71 (3.48 - 8.84) | 1.36 (0.68 - 2.68) | 9.32 (6.40 - 13.10) | 10.37 (7.25 - 14.37) | 1.04 (0.65 - 1.66) | 3.63 (1.92 - 6.21) | 5.32 (3.2 - 8.31) | 1.39 (0.69 - 2.81) |

**Supplementary Table 2.** Incidence, prevalence, mortality and risk ratio of MND in WNSWLHD stratified by sex. Crude rates were calculated using the 2021 Australian Bureau of Statistics census population for WNSWLHD, while age-standardised rates were derived using the Australian population from the 2021 census. Rates are reported for the total population (all-age, 0-85+ years) and for adults aged ≥20 years. 95% confidence intervals are reported within brackets. Risk ratios are calculated as female-to-male.

| Population | Year | Incidence per 100,000 person-years | | | Prevalence per 100,000 population | | | Mortality per 100,000 person-years | | |
|---|---|---|---|---|---|---|---|---|---|---|
| | | All | Male | Female | All | Male | Female | All | Male | Female |
| All-age | 2023 | 4.60 (2.45 - 7.87) | 2.11 (0.44 - 6.18) | 7.11 (3.41 - 13.08) | 8.85 (5.73 - 13.06) | 7.75 (3.87 - 13.86) | 9.96 (5.44 - 16.71) | 1.77 (0.57 - 4.13) | 2.11 (0.44 - 6.18) | 1.42 (0.17 - 5.14) |
| All-age | 2024 | 4.95 (2.71 - 8.31) | 5.64 (2.43 - 11.1) | 4.27 (1.57 - 9.29) | 9.56 (6.30 - 13.90) | 8.45 (4.37 - 14.77) | 10.67 (5.97 - 17.60) | 3.54 (1.7 - 6.51) | 3.52 (1.14 - 8.22) | 3.56 (1.15 - 8.30) |
| All-age | 2025 | 2.48 (1.00 - 5.10) | 2.11 (0.44 - 6.18) | 2.84 (0.78 - 7.28) | 6.02 (3.50 - 9.63) | 7.04 (3.38 - 12.95) | 4.98 (2.00 - 10.26) | 6.02 (3.5 - 9.63) | 3.52 (1.14 - 8.22) | 8.53 (4.41 - 14.91) |
| Populations ≥20 | 2023 | 6.24 (3.32 - 10.67) | 2.90 (0.60 - 8.48) | 9.53 (4.57 - 17.52) | 12.00 (7.77 - 17.72) | 10.64 (5.31 - 19.04) | 13.34 (7.29 - 22.38) | 2.40 (0.78 - 5.6) | 2.90 (0.60 - 8.48) | 1.91 (0.23 - 6.88) |
| Populations ≥20 | 2024 | 6.72 (3.67 - 11.28) | 7.74 (3.34 - 15.25) | 5.72 (2.10 - 12.44) | 12.96 (8.54 - 18.86) | 11.61 (6.00 - 20.28) | 14.29 (8.00 - 23.58) | 4.80 (2.30 - 8.83) | 4.84 (1.57 - 11.29) | 4.76 (1.55 - 11.12) |
| Populations ≥20 | 2025 | 3.36 (1.35 - 6.92) | 2.90 (0.60 - 8.48) | 3.81 (1.04 - 9.76) | 8.16 (4.75 - 13.07) | 9.68 (4.64 - 17.79) | 6.67 (2.68 - 13.74) | 8.16 (4.75 - 13.07) | 4.84 (1.57 - 11.29) | 11.43 (5.91 - 19.97) |

**Supplementary Table 3.** Incidence, prevalence and mortality of MND in WNSWLHD for each year from 2023-2025. Crude rates were calculated using the 2021 Australian Bureau of Statistics census population for WNSWLHD. Rates are reported for the total population (all-age, 0-85+ years) and for adults aged ≥20 years. 95% confidence intervals are included in brackets. Age-standardised rates were not calculated as there were insufficient case numbers for reliable confidence interval calculation.
